# Vitamin D supplementation on colorectal cancer incidence and mortality:a meta-analysis of randomized controlled clinical trials and trial sequential analysis

**DOI:** 10.1101/2022.06.20.22276643

**Authors:** Dou Fu, Wei Yang, Yuanfa Li

## Abstract

**Objective:** To evaluate the effect of oral vitamin D supplementation on colorectal cancer incidence and mortality.

**Methods:** All randomized controlled trials regarding effect of oral vitamin D on colorectal cancer from PubMed, Web of science, Embase and Cochrane Library were searched. Meta analysis and trial sequential analysis of included studies were carried out.

**Results:** Seven RCTs enrolling 72810 participants were included. Compared with placebo, oral vitamin D supplementation was not associated with the incidence of colorectal cancer(RR=1.1,95%CI:0.93-1.31;p=0.28).Subgroup analysis showed no difference in female participants for incidence of colorectal cancer(RR=1.04,95% CI:0.84-1.28;p=0.73). The pooled RR, included 41643 participants, showed no difference in mortality of colorectal cancer(RR= 0.82, 95% CI:0.56 to 1.21; p = 0.32). Trial sequential analysis showed the z-line did not cross the conventional test boundary, TSA monitoring boundary and futility boundaries.

**Conclusion:** Oral vitamin D supplementation offers no benefit on the incidence and mortality of colorectal cancer. However, the RCTs with longer follow-up are needed to ascertain the efficacy of Vitamin D, especially among adults with lower serum 25(OH)D levels in future.

## Introduction

Colorectal cancer(CRC) is one of the most commonly cancers worldwide, and dietary sources with vitamin D(VitD) are considered to be effective for prevention of colorectal cancer [1, 2].The potential beneficial effect of calcium and vitamin D against CRC has also been supported by observational studies[3-6].Additionally, higher serum 25(OH)D was associated with a statistically significant, substantially lower colorectal cancer risk in women[7,8].Although these trials indicated the protective role of oral vitamin D in the risk of CRC, a large clinical trial presented that oral vitamin D did not reduce invasive cancer incidence or cardiovascular events in 2019[9].Interestingly, a meta-analysis and systematic review, including 52 trials in 2019,showed that vitamin D supplementation reduced the risk of cancer death by 16%[10].Whether Vitamin D supplementations reduce the risk of colorectal cancer incidence and mortality remains unclear. Therefore,we conducted the meta-analysis and systematic review to determine the effect of oral vitamin D supplementation on colorectal cancer incidence and mortality.

## Methods

The present meta-analysis was performed in accordance with the recommendations in the Preferred Reporting Items for Systematic Reviews and Meta-Analyses (PRISMA) statement[11].The protocol for this review was registered at PROSPERO (CRD42020209875).

### Search Strategy

The following bibliographical databases were searched: PubMed, Embase, the Cochrane Central Register of Controlled Trials (CENTRAL) and Web of science database dated up to October 1, 2020.The strategies used for searching were including the following terms: (“vitamin D”OR”vitamin D_2_”OR “vitamin D_3_”) and (“Colorectal Cancer”Or “cancer” or “Cancer Incidence” or “mortality”or”Prevention”).

### Study selection and Data Extraction

Randomized controlled trials(RCTs) investigating Vitamin D prevention of colorectal cancer in adults without a history of any cancer were included. Trials were included regardless of date, type of publication or language. Trials assessing non-supplement VitaminD (i.e, food fortified with Vitamin D) were excluded. We also excluded trials using Calcium alone as intervention or trials without data on the outcomes of interest.

Two reviewers(Wei Yang and Dou Fu) independently conducted data extraction and quality assessment, disagreements were resolved by consensus. Data extracted included, in addition to outcomes of interest, information regarding first author, year of publication, country, study design, type and duration of intervention, type of control, numbers of intervention arm and control arm, duration of follow-up, mean age of participants, and gender.

### Assessment for Risk of Bias

Dou Fu and Yuanfa Li evaluated risk of bias (RoB) of all included studies by using Cochrane RoB form version 2.0 independently [12].The RoB2.0 form contains five domains as follows: bias arising from the randomization process, bias due to deviations from the intended interventions, bias due to missing outcome data, bias in measurement of the outcome, and bias in selection of the reported result. The risk of bias in each domain was stratified into “Low risk”, “High risk”or “Some concerns”based on answers to a series of questions. An overall bias was assessed from these five domains.

### Statistical Analysis

Statistical analyses were done with Review Manager 5.4 software (The Cochrane Collaboration). The risk ratio (RR) with 95% confidence interval (CI) and the Mantel-Haenszel method (fixed or random models) were used to analyze dichotomous data. Pooled hazard ratio(HR) was performed by using STATA Version 11.Heterogeneity of the pooled estimates were measured using I^2^ statistics and χ^2^ p-value. Subgroups analysis was performed according to gender, types of VitD(VitD_2_ or VitD_3_),VitD alone or VitD combined with calcium and the duration of follow-up if possible.TSA software (0.9.5.5 Beta, Copenhagen Trial Unit, Copenhagen Trial Unit, Copenhagen, Denmark) was used to perform TSA for the outcomes reported by at least 5 RCTs. The thresholds for the z values were adjusted using O’Brien-Fleming α-spending function to control for type 1 error. The statistical significance was considered when Z curve crossed O’Brien-Fleming α-spending boundaries. The β-spending function and futility boundaries were used for controlling the type 2 error. Crossing the futility boundaries by a Z curve would highlight no difference between the 2 interventions. A 95% confidence level demonstrated the statistical significance. The required information sample(IS) was computed according to 7.5%,10%,15% relative risk reduction(RRR) between two groups and achievement of 80% power. If a sufficient number of trials (ten trials) were found, publication bias was investigated by a funnel plot. The primary outcome was colorectal cancer incidence and the secondary outcome was the mortality from colorectal cancer.

## Results

15830 papers were found from pubmed, Embase, Cochrane Central Register of Controlled Trials (CENTRAL) and web of science.7 papers were finally included after eliminating duplications, reviews, descriptive studies and studies that did not meet the inclusion criteria[9,13-18](Fig.1).

**Figure. 1.**
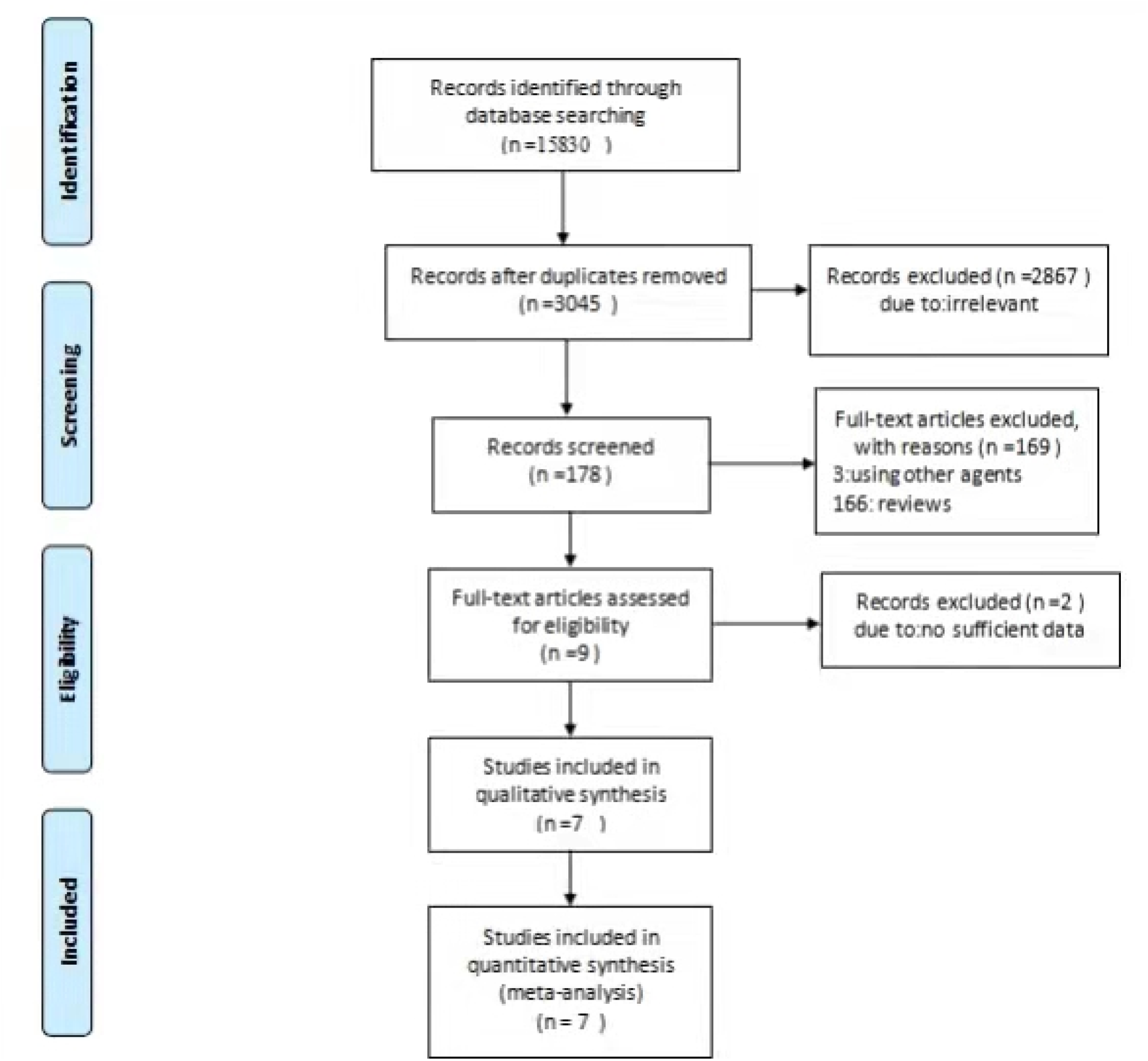
PRISMA Flow Diagram

### Characteristics of included RCTs

7 RCTs were included in the meta-analysis which included 74561 patients(29944 males/44617 females),aging from 45 to79ys.Five RCTs were conducted in the United States, two RCTs in Europe.Vitamin D tables were designed as intervention alone in two trials and Vitamin D tables plus calcium in five trials.The length of follow-up varied from three years to seven years among the studies(Table 1).

**Table 1.**
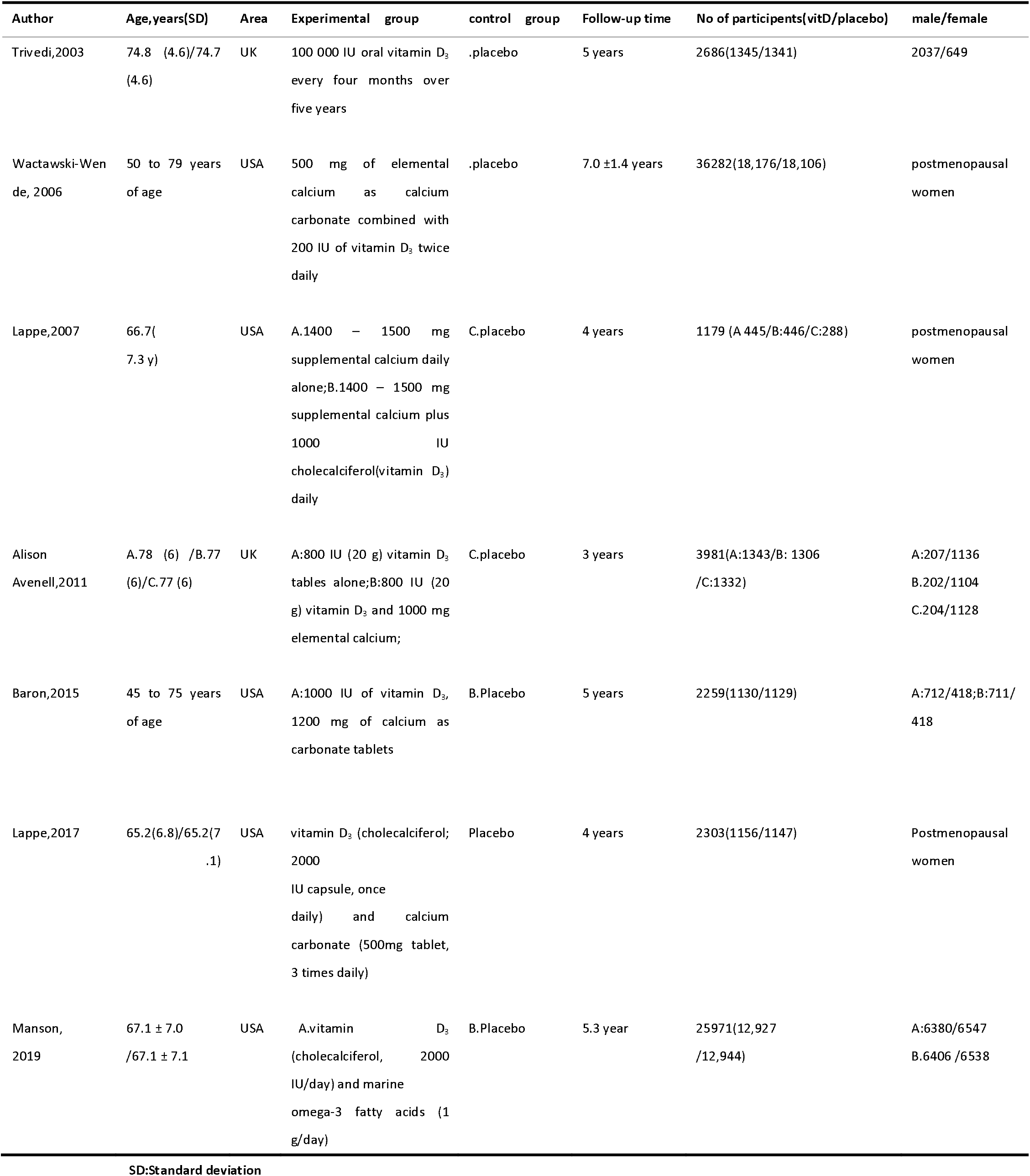
Baseline characteristics of the included studies

### Risk of bias assessment

Allocation concealment was not reported in two trials, and other items were showed. Finally, overall risk of bias was assessed as low risk in five studies except the above two studies(Table 2).

**Table 2.** The risk of bias in included studies

| Study | Bias arising from the randomization process | Bias due to deviations from the intended interventions | Bias due to missing outcome data | Bias in measurement of the outcome | Bias in selection of the reported result | Overall risk of bias |
| --- | --- | --- | --- | --- | --- | --- |
| Trivedi, 2003 | SC | LR | LR | LR | LR | SC |
| Wactawski-Wende 2006 | LR | LR | LR | LR | LR | LR |
| Lappe2007 | LR | LR | LR | LR | LR | LR |
| Alison Avenell 2011 | SC | LR | LR | LR | LR | SC |
| Baron2015 | LR | LR | LR | LR | LR | LR |
| Lappe2017 | LR | LR | LR | LR | LR | LR |
| Manson2019 | LR | LR | LR | LR | LR | LR |

LR: low risk of bias; SC: some concerns

### Incidence of colorectal cancer

7 RCTs, including 72810 patients, reported the incidence of colorectal cancer. There was no difference between two groups (RR=1.1,95% CI,0.93-1.31; p=0.28)(Fig.2). The value for I^2^ suggested a low level of heterogeneity (I^2^ = 0%; p =0.66).Moreover, we also considered the influence of time-to-event outcome and then performed pooled HR. The pooled HR was 1.074 (95%CI:0.877-1.272) without heterogeneity (I^2^ = 0%)(Figure not shown). Finally, sensitivity analysis showed the result kept robust.

**Figure. 2.**
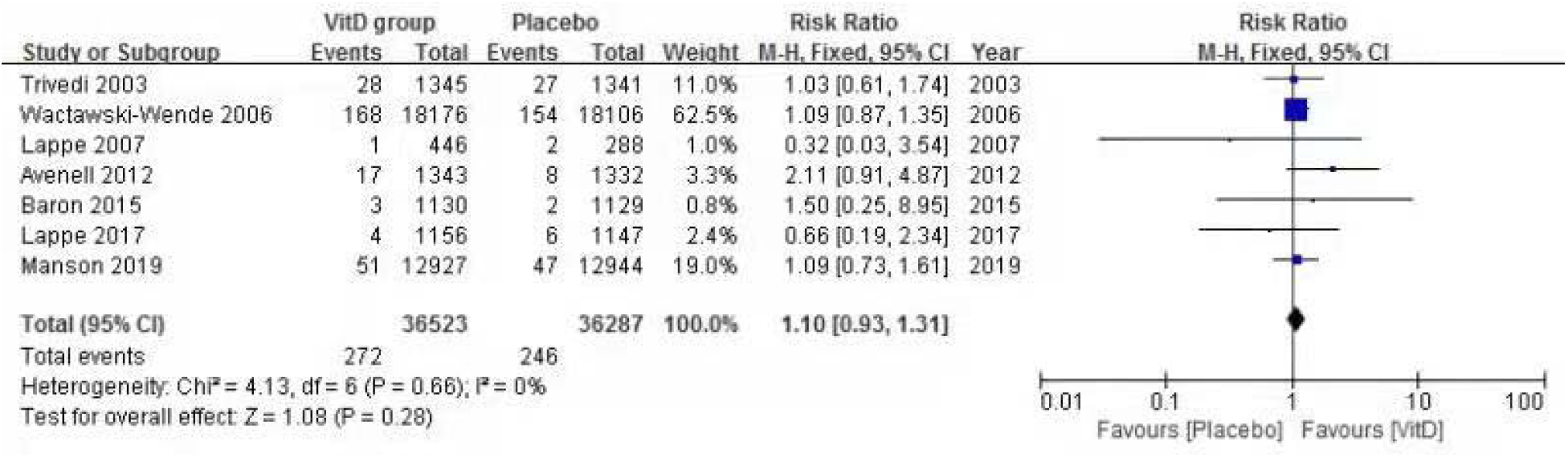
Pooled RR of incidence of colorectal cancer

### Colorectal cancer mortality

Three studies reported death from colorectal cancer were included. Vitamin D supplementation was not superior to placebo on the mortality. (RR= 0.82, 95% CI:0.56 -1.21; p = 0.32)(Fig.3). The value for I^2^ suggested a low level of heterogeneity (I^2^ = 0%; p =0.72). The pooled HR was 0.771 (95%CI:0.436-1.106) without heterogeneity (I^2^=0%).In addition, sensitivity analysis showed the result was robust. Two studies that reported the mortality of females were included, reporting 79 deaths among 36931 female patients. Vitamin D supplementation was not superior to placebo related to mortality in females(OR= 0.75, 95% CI:0.48 to 1.18; p = 0.21)(Fig.4). The value for I^2^ suggested a low level of heterogeneity (I^2^ = 45%; *p* =0.18).

**Figure. 3.**
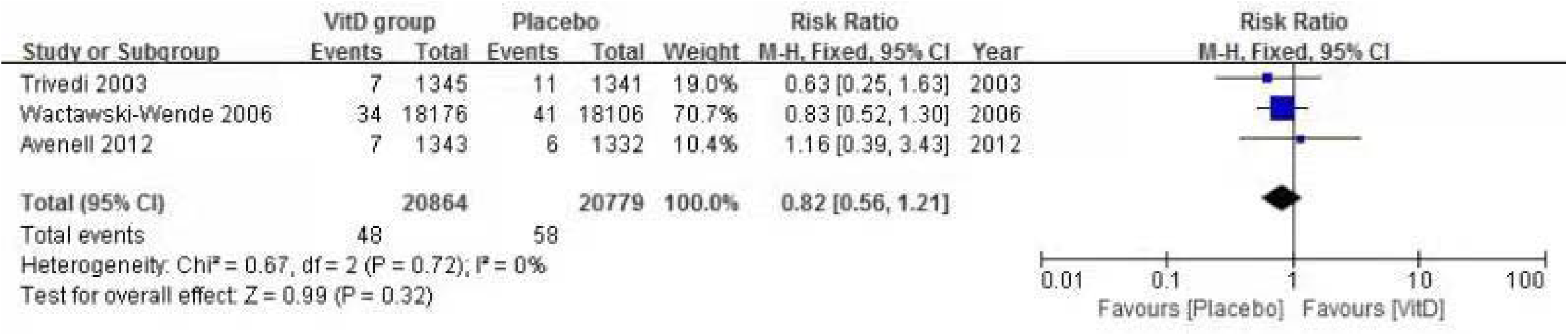
Pooled RR of mortality of colorectal cancer

**Figure. 4.**
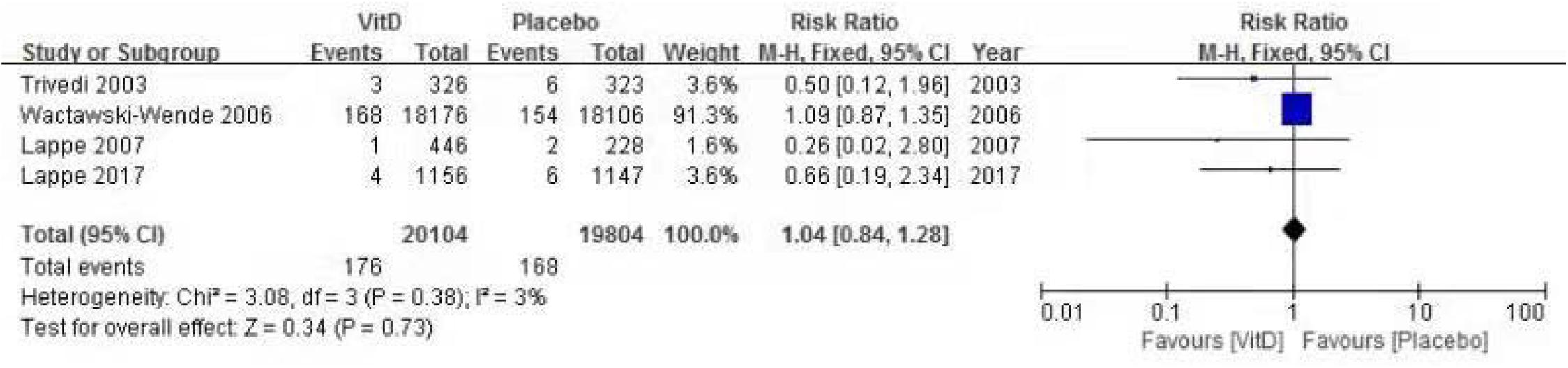
Pooled RR of mortality of colorectal cancer in females

### Subgroup analysis and publication bias

Subgroup analysis was performed to reveal the influence of gender, the length of follow-up and the intervention(VitD alone or VitD plus calcium) on the colon cancer incidence. The three subgroups showed that no significant differences were found between the two groups(Table 3).

**Table 3.** Subgroup analysis

| Subgroups title | No of studies | No of patients | RR,95%CI | Heterogeneity |
| --- | --- | --- | --- | --- |
| **Sex** |  |  |  |  |
| female | 3 | 39319 | 1.06(0.86,1.31) | I<sup>2</sup>=0%,P=0.46 |
| Male and female | 4 | 33491 | 1.18(0.88,1.57) | I<sup>2</sup>=0%,P=0.51 |
| **Follow-up time** |  |  |  |  |
| ≥5years | 4 | 67098 | 1.08(0.91,1.30) | I<sup>2</sup>=0%,P=0.98 |
| <5 years | 3 | 5712 | 1.32(0.70,2.48) | I<sup>2</sup>=46%,P=0.16 |
| **Intervention** |  |  |  |  |
| Vitamin D alone | 2 | 31232 | 1.17 ( 0.87,1.57 ) | I<sup>2</sup>=11%,P=0.32 |
| Both vitamin D and calcium | 5 | 41578 | 1.06 ( 0.86,1.32 ) | I<sup>2</sup>=0%,P=0.64 |

RR:risk ratio;95%CI:95%confidence interval

Due to the number of included trials was not above ten, publication bias was not investigated by a funnel plot.

### Trial sequential analysis

We conducted TSA on colon cancer incidence (see supplementary figure1,2,3). A RRR of 7.5%, 10%, 15% with alpha error (two sided) of 5% and power of 80% was estimated. When RRR was7.5%, 10% or 15%, the cumulative z-line did not cross the conventional test boundary and the trial sequential monitoring boundary. But The cumulative z-line did also not cross the futility boundaries.

## Discussion

In this meta-analysis, vitamin D supplementation had no detectable effect on the incidence and mortality of colorectal cancer. The intervention also did not reduce the incidence and mortality of colorectal cancer across subgroups, including postmenopausal women or males and females, vitamin D intake alone or vitamin D combined with calcium and different length of follow-up.

Although TSA reflected that the cumulative z-line did not cross the futility boundaries when RRR varied from 7.5% to 15%,the result seemed to show a trend that does not support the effectiveness of the supplementation on the prevention of colorectal cancer. The pooled HR, not reported previously, also was consistent with RR.

To our knowledge, the study was the first meta-analysis focusing on the potential chemopreventive effect of Vit D supplementation on colorectal cancer. Yu Zhang and colleagues [10] found that the risk ratio (RR) for risk of cancer mortality was 0.84 (95% CI: 0. 74–0.95), and represented that oral vitamin D reduced the risk of cancer death by 16%. Similarly, Keum et al conducted an updated meta-analysis of RCTs, which also demonstrated a beneficial effect of vitamin D supplementation on risk of total cancer mortality and no benefit on total cancer incidence[19].Both of these previous meta-analyses included total cancer, whereas only colorectal cancer was concerned in our study. Though the two meta-analyses found the potential benefit of vitamin D on total cancer mortality, the true effect may be hided among other solid cancer but not colorectal cancer. Across the included trials in our study, one of three trials which showed the result of death due to colorectal cancer, reported a lower risk of mortality of colorectal cancer among women in VitD group (p=0.04)[13].But pooled RR and 95%CI in this study showed no significant difference in female participants or both gender participants. Compared to total cancer incidence and mortality during follow-up, the mortality of colorectal cancer was lower, which may be not reflected statistically. In this sense, detection of reduced mortality because of colorectal cancer, if present, may require more participants.

Here, we also need to raise two issues as follows:1)serum 25-hydroxyvitamin D(25(OH)D) and risk of colorectal cancer;2)the length of follow-up and the effect of VitD.

Previous basic studies demonstrated that VitD, by binding with VDR, can induce apoptosis of colon cancer cells and counteract aberrant Wnt/β-catenin signaling and then, inhibit proliferation of human colon cancer cells[20,21].Meanwhile,in observational studies,serum 25(OH)D concentration,75nmol/L-100nmol/L,was related to a 19%-27% reduction of colorectal cancer risk[7].And,meta-analyses showed an inverse association with lower risk of colon cancer in persons with higher 25(OH)D levels and revealed an association between higher blood 25(OH)D concentrations and better survival in CRC patients [22-24]. These findings above seemed to favour the hypothesis of protective effect of vitamin D from dietary intake with/without supplementation against the risk of colorectal cancer.

Interestingly, among the included trials in our study,four trials presented mean serum total 25(OH)D level at baseline and after 1 to 4 years. Mean serum 25(OH)D level concentrations, increased to 41.8ng/mL-45.1 ng/mL or 96nmol/L,were higher in the VitD group after treatment than placebo group [9,13,15,18].But,no positive findings that supported VitD supplementation as a protective agent against colorectal cancer were found.Meanwhile,calcium and vitamin D supplementation designed in RCTs was also found to have no benefit on recurrent colorectal adenomas and even contrarily increased the risk of colorectal cancer precursors such as sessile serrated adenomas or polyps[17,25].The protective effect of vitamin D,concluded by observational studies,was not supported by RCTs.There may be,we thought,some reasons why the results of cohort studies were not consistent with RCTs.Firstly,the nature of observational studies is different with RCTs. Confounding factors(i.e,multvitamin from food,age,gender) in observational studies may result into bias even if the manager controlled them as mostly as possible.Secondly,mean serum 25(OH)D level at baseline were balanced in RCTs but not in observational studies.Finally, the follow-up was likely shorter (3-7years,mostly below 5 years) in RCTs than in observational studies(1-15years,mostly above 5years). Therefore, we speculated that a potential protective effect of VitD on the risk of colorectal cancer may exist among human beings with lower serum 25-hydroxyvitamin D levels but not in normal VitD status people.

As we know, most solid tumors take some decades to develop in adults. If the protective effect of VitD with or without calcium supplementation against colorectal cancer will be reflected at around 10 years after intervention, the follow-up of 3-7 years in our study or included RCTs may have been not sufficient to present an effect. Although we did not find a trend toward protection in the study, we could not ignore a relationship between vitamin D intake/status and the risk of CRC, which was demonstrated by many observational and basic studies. Therefore, due to the long latency for colorectal cancer development, longer follow-up is necessary to fully detect potential protective effect.

There were some limitations in this study. Firstly, the number of included trials was insufficient so that we could not perform the subgroups according to different oral time and dose of vitamin D. Publication bias was also not conducted. Secondly, different oral time and dose of vitamin D across the included trials may result into risk of bias. Thirdly, all of the included participants were older adults, whereas the data regarding younger adults was lacking. All these limitations might lead to bias of the clinical efficacy.

In conclusion, although the quality of most included trials among the meta-analysis was high, our results did not support the protective effect of VitD against the incidence and mortality of colorectal cancer. Based on the current evidence, VitD supplementation is not suitable for the prevention of colorectal cancer. However, the RCTs with longer follow-up are needed to ascertain protective effect of VitD, especially among vitamin D deficiency adults.

## Data Availability

All data produced in the present work are contained in the manuscript

---

**Supplementary figure 1.**
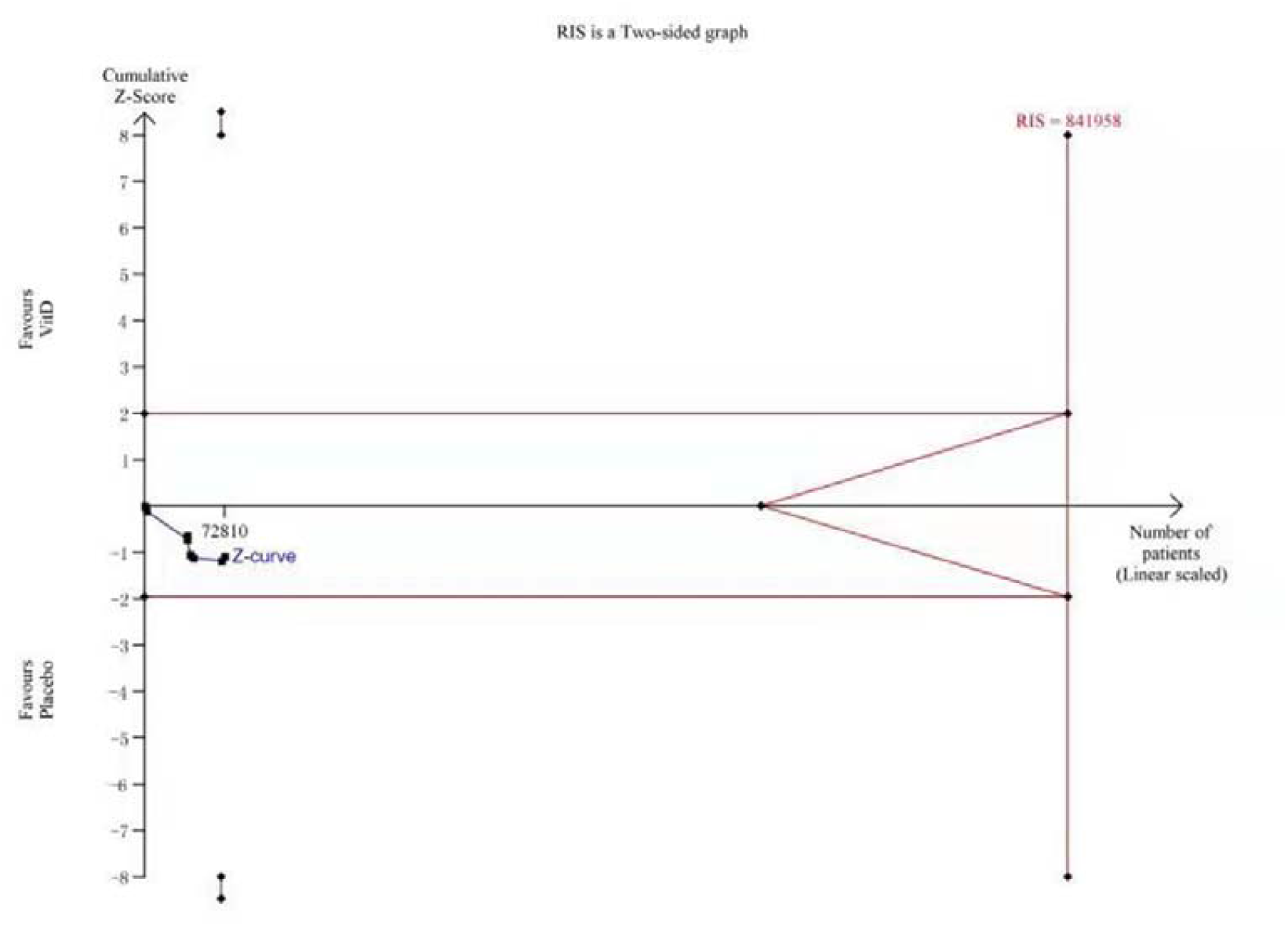
A diversity-adjusted information size (DIS) size of 841,958 patients was calculated based on an anticipated relative risk reduction (RRR) of 7.5% (α=0.05 (two-sided), β=0.20). The blue cumulative z-curve was constructed using a fixed-effects model and not crossed any boundary

**Supplementary figure 2.**
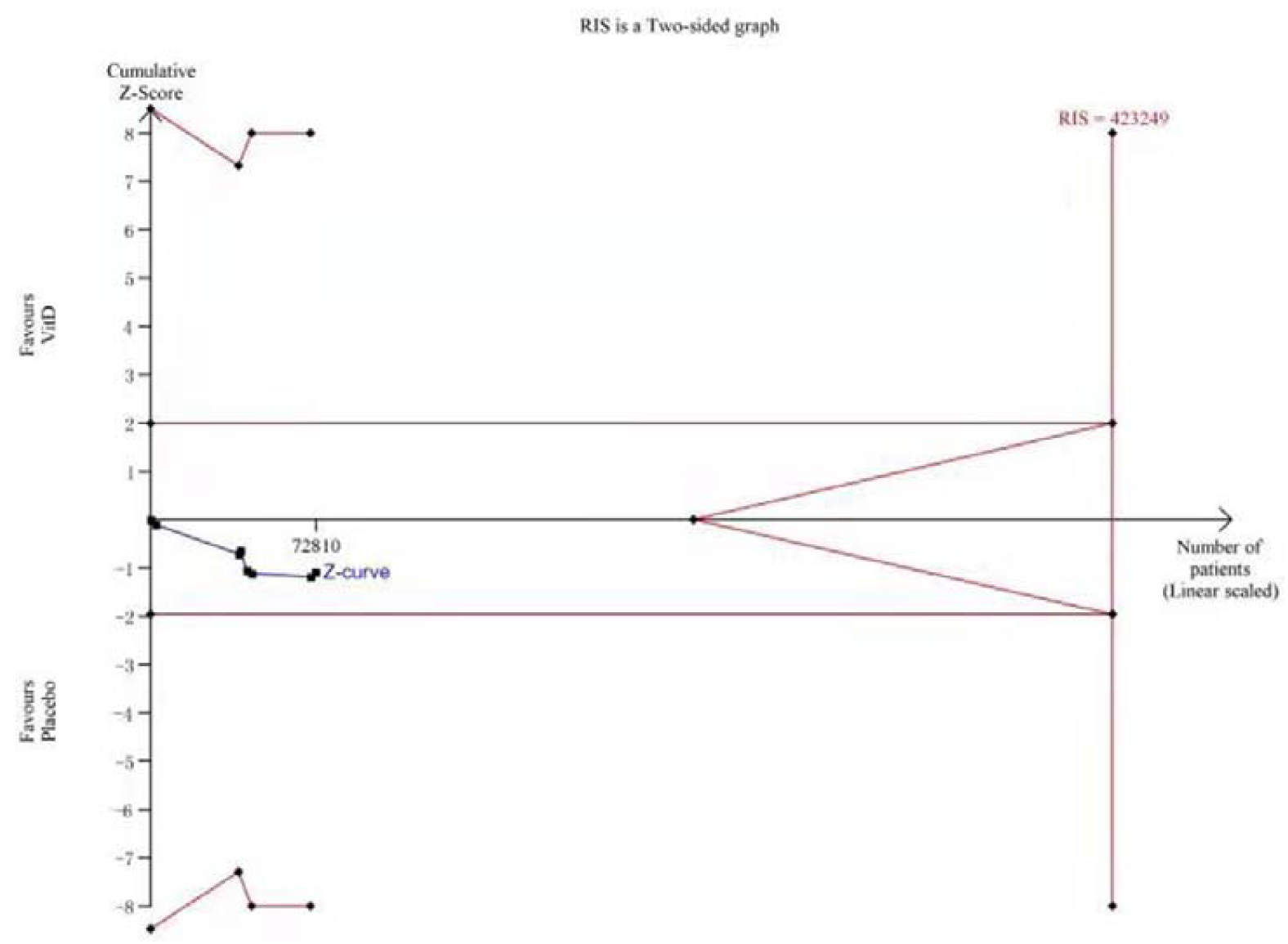
A diversity-adjusted information size (DIS) size of 423,249 patients was calculated based on an anticipated relative risk reduction (RRR) of 10% (α=0.05 (two-sided), β=0.20). The blue cumulative z-curve was constructed using a fixed-effects model and not crossed any boundary.

**Supplementary figure 3.**
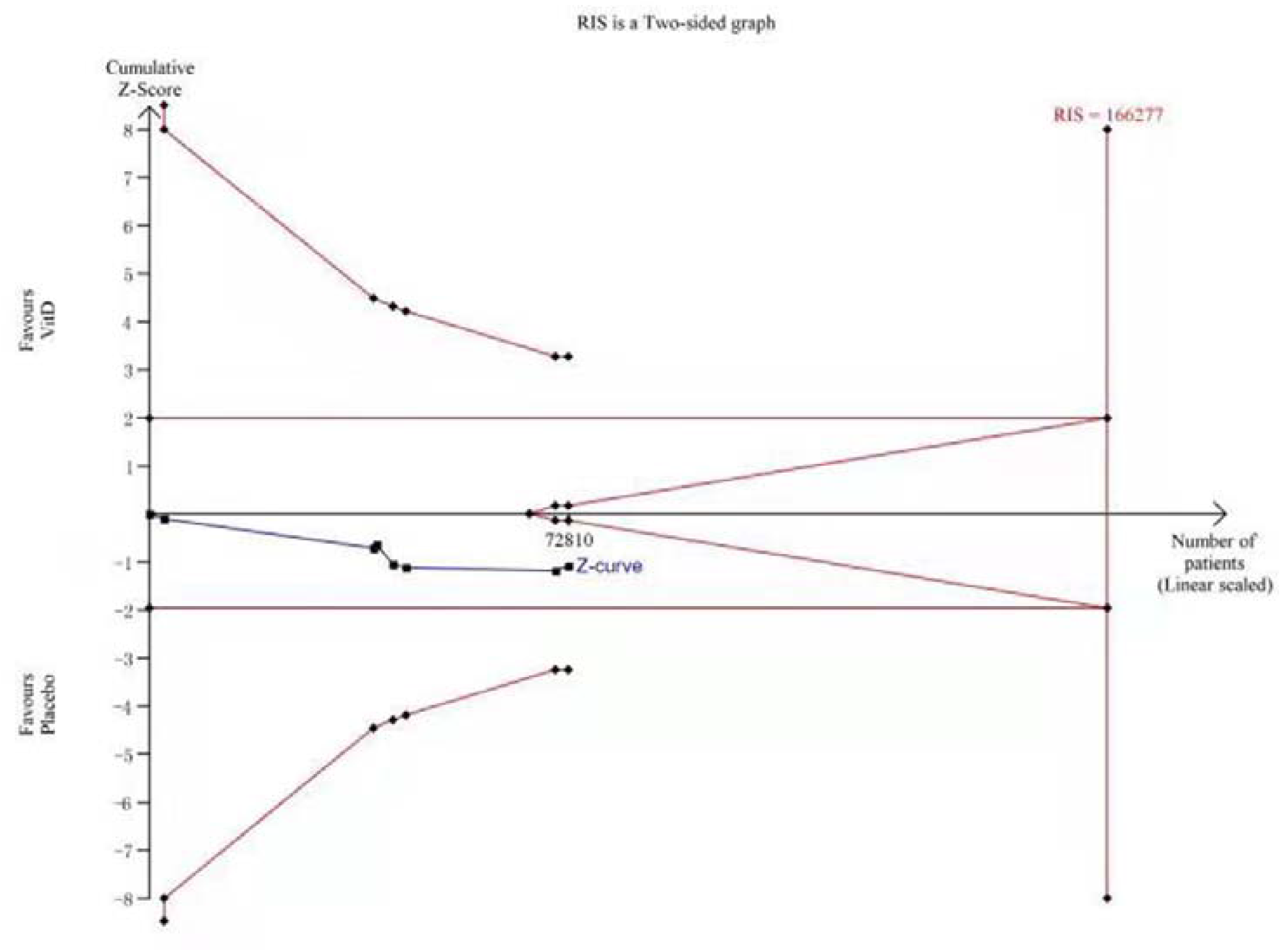
A diversity-adjusted information size (DIS) size of 166,277 patients was calculated based on an anticipated relative risk reduction (RRR) of 15% (α=0.05 (two-sided), β=0.20). The blue cumulative z-curve was constructed using a fixed-effects model and tended to cross the boundary for futility.

